# Nightshift Imposes Irregular Lifestyle Behaviors in Police Academy Trainees

**DOI:** 10.1101/2023.07.07.23292363

**Authors:** Melissa L. Erickson, Rebecca North, Julie Counts, Will Wang, Kathryn N. Porter Starr, Laurie Wideman, Carl Pieper, Jessilyn Dunn, William E. Kraus

**Affiliations:** Translational Research Institute, AdventHealth; Center for the Study of Aging and Human Development, Duke University; Duke Molecular Physiology Institute, Duke University; Department of Biomedical Engineering, Duke University; Geriatric Research, Education, Clinical Center, Durham VA Health Care System; Department of Kinesiology, University of North Carolina at Greensboro; Department of Biostatistics and Bioinformatics, Duke University

## Abstract

**Study Objective:** Shiftwork increases risk for numerous chronic diseases, which is hypothesized to be linked to disruption of circadian timing of lifestyle behaviors. However, empirical data on timing of lifestyle behaviors in real-world shift workers are lacking. To address this, we characterized the regularity of timing of lifestyle behaviors in shift-working police trainees.

**Methods:** Using a two-group observational study design (N=18), we compared lifestyle behavior timing during 6 weeks of in-class training during dayshift, followed by 6 weeks of field-based training during *either* dayshift or nightshift. Lifestyle behavior timing, including sleep/wake patterns, physical activity, and meals, was captured using wearable activity trackers and mobile devices. The regularity of lifestyle behavior timing was quantified as an index score, which reflects day-to-day stability on a 24h time scale: Sleep Regularity Index (SRI), Physical Activity Regularity Index (PARI) and Mealtime Regularity Index (MRI). Logistic regression was applied to these indices to develop a composite score, termed the Behavior Regularity Index (BRI).

**Results:** Transitioning from dayshift to nightshift significantly worsened the BRI, relative to maintaining a dayshift schedule. Specifically, nightshift led to more irregular sleep/wake timing and meal timing; physical activity timing was not impacted. In contrast, maintaining a dayshift schedule did not impact regularity indices.

**Conclusion:** Nightshift imposed irregular timing of lifestyle behaviors, which is consistent with the hypothesis that circadian disruption contributes to chronic disease risk in shift workers. How to mitigate the negative impact of shiftwork on human health as mediated by irregular timing of sleep/wake patterns and meals deserves exploration.

## INTRODUCTION

Maintaining regular circadian rhythms is an important feature of health ^1–3^. Behavior and endogenous circadian clocks have a bi-directional relationship; this premise underlies the concept that maintaining optimal behavioral patterns promotes robust circadian rhythmicity and mitigates disease ^4^. In contrast, a lifestyle characterized by irregular behavior patterns— particularly in which behaviors are misaligned with endogenous circadian rhythms—may increase susceptibility to chronic disease. In support, epidemiological evidence indicate shiftwork is associated with numerous chronic diseases ^5–9^. These findings suggest that discordance between behavioral timing and intrinsic circadian timing contributes to negative health consequences.

Previous studies have demonstrated the negative consequences of circadian disruption using highly controlled inpatient studies, and these experimental models imply that nightshift work leads to acute disruptions in health ^10–12^. However, the extent to which these findings translate to settings of real-world shift workers is currently unknown. Without this information, it is difficult to predict the extent to which treatment interventions focusing on maintaining circadian rhythms would be efficacious for disease prevention and treatment specific to shift workers. To partly address this knowledge gap, the human circadian field needs more tools that quantify behavior regularity in field-based settings. Wearable technology and smart phone devices offer promise here, enabling real-world data capture applicable over long durations and scaled up for assessment of human behaviors that are irregular or misaligned during shiftwork, with all of their subsequent downstream negative health consequences ^13–15^.

The next step towards developing a metric that quantifies behavior regularity is determining analytical approaches that can be readily applied to data collected in field settings. One previous approach developed an index scoring method to quantify the regularity of sleep/wake cycles. Termed the sleep regularity index (SRI), this metric quantifies the likelihood of sleep/wake episodes occurring at the same time within a 24h time scale, or day-to-day variation ^16, 17^. In addition, mealtime regularity has been previously quantified by a metric termed Composite Phase Deviation ^18, 19^. Informed by these previous approaches, we sought to expand and combine these metrics to integrate a composite score accounting for three behavior patterns that are known to impact health status: sleep/wake patterns, physical activity, and meals.

The primary objective of this study was to characterize the effects of nightshift on the regularity of lifestyle behavior timing in a real-world setting. We used wearable activity trackers to assess sleep/wake patterns and physical activity patterns, and mobile devices to assess meal timing. We conducted these assessments in two groups of police trainees that followed a real-world shiftwork schedule: one group that transitioned from dayshift to nightshift, versus a comparator group that maintained a dayshift schedule. Accordingly, we assessed day-to-day behavior regularity for three behaviors: sleep/wake patterns, physical activity, and meals. We then combined these indices into a composite score, termed the Behavior Regularity Index (BRI). We hypothesized that nightshift, incorporating a cyclic pattern of night work and off days, would be characterized by irregular behavior patterns. To test this hypothesis, we compared changes in lifestyle behavioral regularity (as measured by the BRI) during the transition from a dayshift to nightshift schedule relative to changes in lifestyle behavioral regularity (BRI) during maintenance of a dayshift schedule.

## METHODS

### Study Design

The study design has been previously described ^13^. In brief, this was a two-group observational repeated measures study design. Police trainees followed 24 weeks of in-class training and 14 weeks of field-training, and the current study protocol was a short-term observational period nested within the training schedule. Specifically, we assessed behaviors during the last six weeks of in-class training (baseline) and the first six weeks of field training, totaling 12 weeks of observation.

In-class training involved classes held Monday to Friday during daytime hours (7:30 AM-5:00 PM). Field-training involved either a day or night shiftwork schedule. Day shifts included Shift A: 6 AM-5 PM and Shift B: 10 AM to 9 PM, whereas night shifts included Shift C: 4 PM-3 AM and Shift D: 8 PM-7 AM. The same shift (A, B, C, or D) was maintained throughout the 6-week observation period, with trainees operating on a 4-day on, 4-day off schedule. No distinction was made in the analysis between weekdays and weekends during in-class training or days on/off during shiftwork so as to capture the level of irregularity in the trainees’ behaviors, whether imposed or voluntary.

#### Participants

Study inclusion criteria have been previously described ^13^. These included: 1) being enrolled in a local public safety training program; and 2) owning a smartphone. Institutional Review Board approval was given by Duke University Health System Institutional Review Board for Clinical Investigations (IRB# Pro00077319). All participants gave informed consent prior to participation in this study.

### Study Protocol

As described above, this was a field-based study. During the 12-week protocol, participants wore activity trackers (Garmin vívosmart® HR, Olathe, KS) and used smart phones to report recalled mealtimes. From these collection methods, we determined behavioral variability for the following three behaviors: sleep/wake patterns, physical activity, and meals, which were then used to calculate the BRI.

#### Wearable Assessments

Activity trackers were used to assess sleep/wake patterns and physical activity during in-class training and field-based training. The Garmin vívosmart® HR was worn on the wrist 24/7 (except when charging device) ^13^. This activity tracker provides readouts of “activity level” and heart rate, which are used to inform sleep/wake labeling in 15-minute epochs. Garmin vívosmart® HR wear time of 80% over 24h was the criteria used to determine data completeness. To be considered for data analysis, at least 50% of days were required to meet wear time criteria. We applied a novel sleep-labeling method to improve sleep/wake labeling completeness ^13^. This method involves a post-data processing step to increase labeling accuracy of daytime sleep episodes – which we previously observed were frequently mislabeled as non-sleep among nightshift workers. We used this sleep-labeling method to determine sleep initiation time and sleep duration.

The Garmin vívosmart® HR was also used to determined exercise/physical activity duration and intensity. For this, we used the Garmin mean motion intensity score. This is a proprietary method that takes acceleration and heart rate into account to average motion intensity levels within 15-minute epochs, resulting in values ranging from 0 to 7.

#### Meal Timing Assessments

Self-reported mealtimes were collected throughout the duration of the study protocol. Starting the day after enrollment, participants responded to a text message prompt that inquired about the times of their major meals during the prior day. This text message prompt was sent at 4 PM regardless of shiftwork schedule, and participants replied when it was convenient for them to do so.

### Regularity Indices

The behaviors of interest – sleep/wake, physical activity, and meal – were separately quantified using the following 3 measures: Sleep Regularity Index ^16, 17^, Physical Activity Regularity Index, and Mealtime Regularity Index. Each of these metrics range in value from 0 to 100 and are further described below.

The *Sleep Regularity Index (SRI)*, in which sleep vs. awake is assessed using our sleep detection algorithm in 15-minute epochs, has been described by others ^16, 17^ but is reiterated here for completeness. For a given subject, the SRI is calculated across the *J* 15-minute epochs in *K* observed days as:

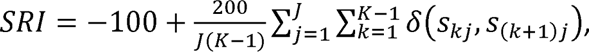

where *s_kj_* denotes the subject’s sleep/wake status (0 or 1) on day *k* during the *j*th 15-minute epoch, and 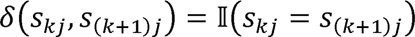 is an indicator function that assigns the value 1 if the subject has the same status, asleep or awake, during the *j*th epoch on consecutive days, or 0 otherwise. Although the SRI could theoretically take values between [-100,100], the developers noted that negative values are highly unlikely to be observed and so the practical range of values is [0,100] ^16^.

The *Physical Activity Regularity Index (PARI)*, in which activity is binned within 15-minute epochs as low [0-3], medium [4-6], or high [7] by rounding mean motion intensity scores (from Garmin), is computed similarly as the previously described SRI. Specifically, for a given subject, the PARI is calculated across the *J* 15-minute epochs in *K* observed days as:

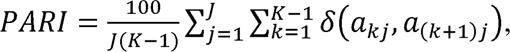

where *α_kj_* denotes the participant’s physical activity level (low, medium, or high) on day *k* in epoch *j*, and δ(*a_kj_*, *a_(k+l)j_*) is the indicator function comparing activity levels during the same 15-minute epoch across consecutive days. The PARI is scaled to take values in [0,100] by multiplying the average similarity index by 100. This difference in scaling, relative to the SRI, is due to the increased variability associated with three possible physical activity levels rather than two, as for sleep. Scaling the PARI as the SRI would result in negative PARI values, which are not easily interpretable.

The *Mealtime Regularity Index (MRI)* assesses day-to-day stability of meal timing. The MRI extends the metric Composite Phase Deviation (CPD), which was initially used to quantify circadian misalignment in sleep patterns ^19^ and later applied to mealtimes ^18^. CPD measures variability by combining how different meal timing is compared to that on the previous day (regularity) and how far away meal timing is from the average mealtime (alignment). For a given participant and meal, CPD is calculated across days *k* = 1, …, *K* as follows:

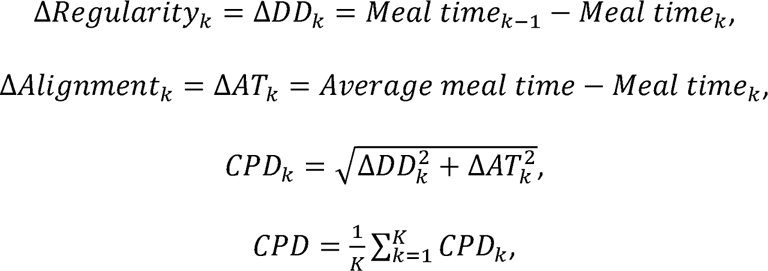

To scale the index value between [0,100] and invert it to indicate regularity, we let 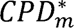 = (1 - [*CPD_m_*/*CPD_max_*]) * 100, where *CPD_max_* assumes meal m ε {1,3} alternates timing by 12 hours each day yielding Δ*DD* = 12, Δ*AT*= 6, and *CPD_max_* = 6√5. The MRI, which reflects a participant’s average regularity across meals, in hours, relative to a perfectly regular pattern of meal timing, can then be defined as

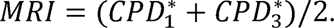

Using the 3 indices described above, we developed the *Behavior Regularity Index (BRI)*. For a given participant, the BRI is the predicted response from a multiple logistic regression model with the SRI, PARI, and MRI as predictors and expected regularity status as the response *Y*, as given below:

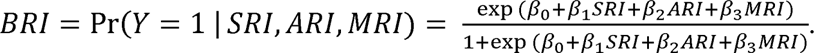

Here, the expected regularity status corresponds to whether the trainee was following a dayshift (1) or nightshift (0) schedule, regardless of actual observed behavior, with the *a priori* assumption that nightshift workers revert to a typical daytime schedule on days not working. Succinctly, the BRI measures the degree, ranging from 0 to 1, to which a participant is performing behaviors in a regular pattern, day-to-day. A BRI value of 0 indicates complete irregularity and 1 indicates perfect regularity.

Each component index was calculated separately for in-class training and field-based training. For the MRI, average mealtime was determined during in-class training as a circular mean (i.e., in polar coordinates then converted back to hours) and used for both in-class and field-based training computations. The logistic regression model producing the BRI was then trained on the component indices from the two time periods (in-class training and field-based training), where each trainee thus contributed two observations. We assumed independence of observations within subjects because accounting for paired samples with a grouping covariate resulted in overfitting and the N-1 grouping coefficients cannot be applied in predictions of new observations.

### Statistical Analysis

R version 4.2.1 was used for mathematical computation and data visualization. Statistical analyses were conducted with SAS software, Version 9.4 (SAS Institute Inc., Cary, NC, USA). We first determined the impact of transitioning from in-class training to field-based training in both the dayshift and nightshift group (within-group differences) in the SRI, PARI, MRI, and the composite BRI using the Wilcoxon signed rank test. To assess the impact of nightshift on behavior regularity, we next determined if the in-class to field-training transition was different between the dayshift and nightshift groups (between-group differences) on the same indices using the Kruskal-Wallis test of equality test. Behavior regularity during in-class training was compared between the dayshift and nightshift groups using the Wilcoxon rank sum test. Regularity indices are summarized and reported as median (Q1, Q3). Significance of within- and between-group differences was accepted at *P* < 0.05. Estimated odds ratios, 95% confidence intervals, and the corresponding *P*-values are also reported for each of the three component indices of the BRI.

## RESULTS

### Participant Characteristics

The current study cohort consisted of 18 individuals, which is a subset of our previous cohort ^13^. Participants lacking meal timing data or who were not assigned to one of the four shiftwork schedules were excluded. The mean age was 26 (± 6.7) yr, and mean BMI was 26 (± 3.8) kg/m^2^. Representative polar plots display the impact of transitioning from a dayshift to a nightshift schedule on sleep-wake patterns, physical activity, and mealtimes, relative to maintaining a dayshift schedule (**Figure 1**).

**Figure 1:**
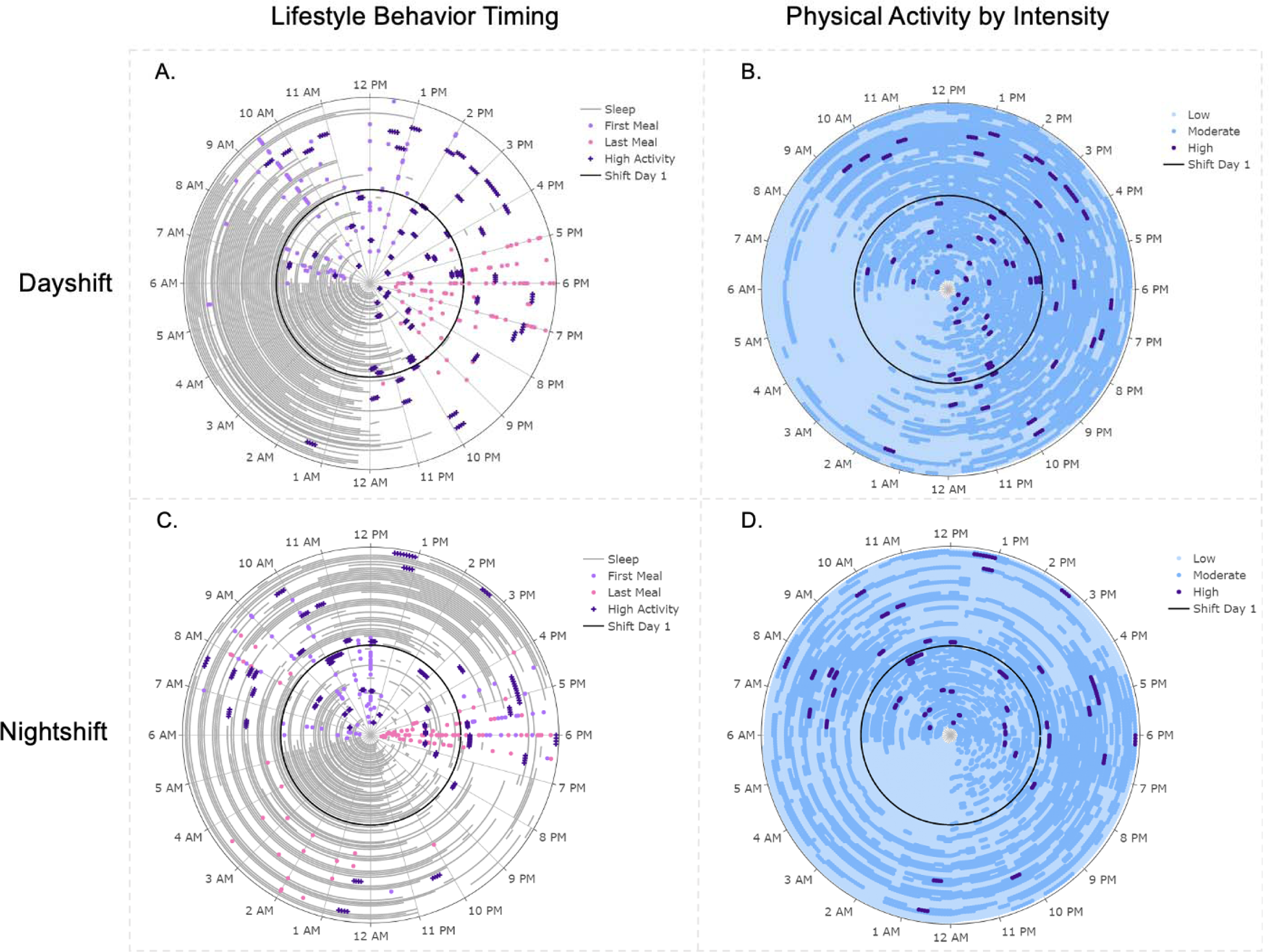
Lifestyle Behavior Timing on Dayshift versus Nightshift. **Panels A and B** show representative polar plots for an individual participant during 6 weeks of dayshift in-class training followed by 6 weeks dayshift field-based training. **Panels C and D** show representative polar plots for an individual participant during 6 weeks of dayshift in-class training followed by 6 weeks nightshift field-based training. Panels A and C show the occurrence of sleep periods (grey lines), meals (light purple and pink), and physical activity bouts (dark purple). Panels B and D show the occurrence of physical activity bouts by intensity, including low (light blue), moderate (blue), and high (dark purple). The transition from in-class training to field-based training at 6 weeks is indicated by the black concentric circle.

### Effect of Shiftwork on Behavior Regularity: Input Indices

#### Sleep Regularity Index

We have reported the SRI for the larger participant cohort elsewhere^13^. Transitioning from dayshift in-class training to dayshift field-based training did not significantly reduce the SRI (70.3 vs. 64.9; *P*=0.578; **Table 1**), whereas transitioning from dayshift in-class training to nightshift field-based training significantly reduced the SRI (56.8 vs. 46.6; *P*=0.042; **Table 1**). However, the changes in the SRI were not different between groups (*P*=0.342; **Table 1**). No differences were observed between groups during the in-class training dayshift periods (*P*=0.147). **Figure 2** shows density plots of the SRI during all four conditions.

**Figure 2:**
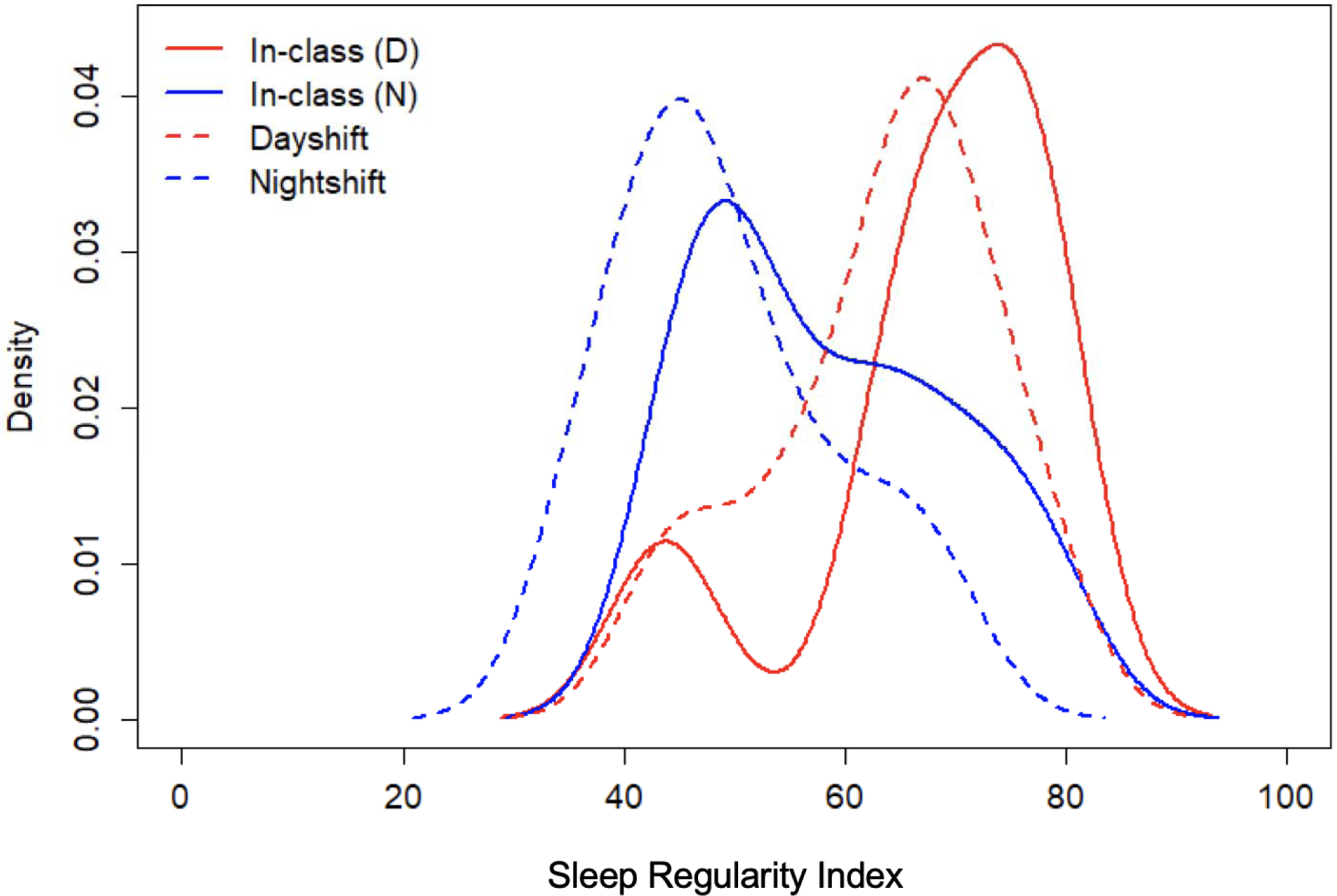
Regularity of Sleep/Wake Timing is Reduced on Nightshift. Figure shows density plots of Sleep Regularity Index (SRI) of the participant cohort during the transition from dayshift in-class training (In-class (D); red line) to dayshift field-training (Dayshift; dashed red line), as well as during the transition from dayshift in-class training (In-class (N); blue line) to nightshift field-training (Nightshift; dashed blue line). Each index is expressed as a value ranging from 0 to 100, in which 0 reflects irregularity and 100 reflects regularity.

**Table 1:** Lifestyle Behavior Regularity Indices on Dayshift and Nightshift

| **Indices** | **<u>Dayshift</u>** |  |  | **<u>Nightshift</u>** |  |  | **Between-Groups**<br>***P*-value** |
| --- | --- | --- | --- | --- | --- | --- | --- |
|  | In-Class | Field-Based | Within-Groups<br>*P*-value | In-Class | Field-Based | Within-Groups<br>*P*-value |  |
| Sleep Regularity | 70.3 (64.6, 77.1) | 64.9 (55.1, 69.9) | 0.578 | 56.8 (48.2, 69.0) | 46.6 (42.6, 56.7) | 0.042* | 0.342 |
| Physical Activity Regularity | 63.5 (61.2, 67.7) | 63.4 (56.6, 67.8) | 0.297 | 60.9 (56.7, 65.5) | 59.0 (52.7, 62.1) | 0.365 | 0.751 |
| Mealtimes Regularity | 84.5 (82.4, 88.5) | 84.1 (76.8, 91.0) | 0.375 | 85.5 (83.6, 86.5) | 49.2 (35.3, 74.0) | 0.001* | < 0.001* |
| Behavior Regularity | 1.00 (1.00, 1.00) | 1.00 (0.82, 1.00) | 0.250 | 1.00 (1.00, 1.00) | 0.00 (0.00, 0.24) | 0.001* | < 0.001* |

Values presented as median (Q1, Q3). \* indicates $P \leq 0.05$ .
Within-Groups *P*-values from Wilcoxon signed rank tests applied to within-group differences.
Between-Groups *P*-values from Kruskal-Wallis test of equality of within-group differences.

#### Physical Activity Regularity Index

The PARI was not significantly impacted by either transitioning from dayshift in-class training to dayshift field-based training (63.5 vs. 63.4, *P*=0.297; **Table 1**) or transitioning from dayshift in-class training to nightshift field-based training (60.9 vs. 59.0, *P*=0.365; **Table 1**). Thus, changes in the PARI were not different between groups (*P*=0.751; **Table 1**). No differences were observed between groups during the in-class training dayshift periods (*P*=0.085). **Figure 3** shows density plots of the PARI during all four conditions.

**Figure 3:**
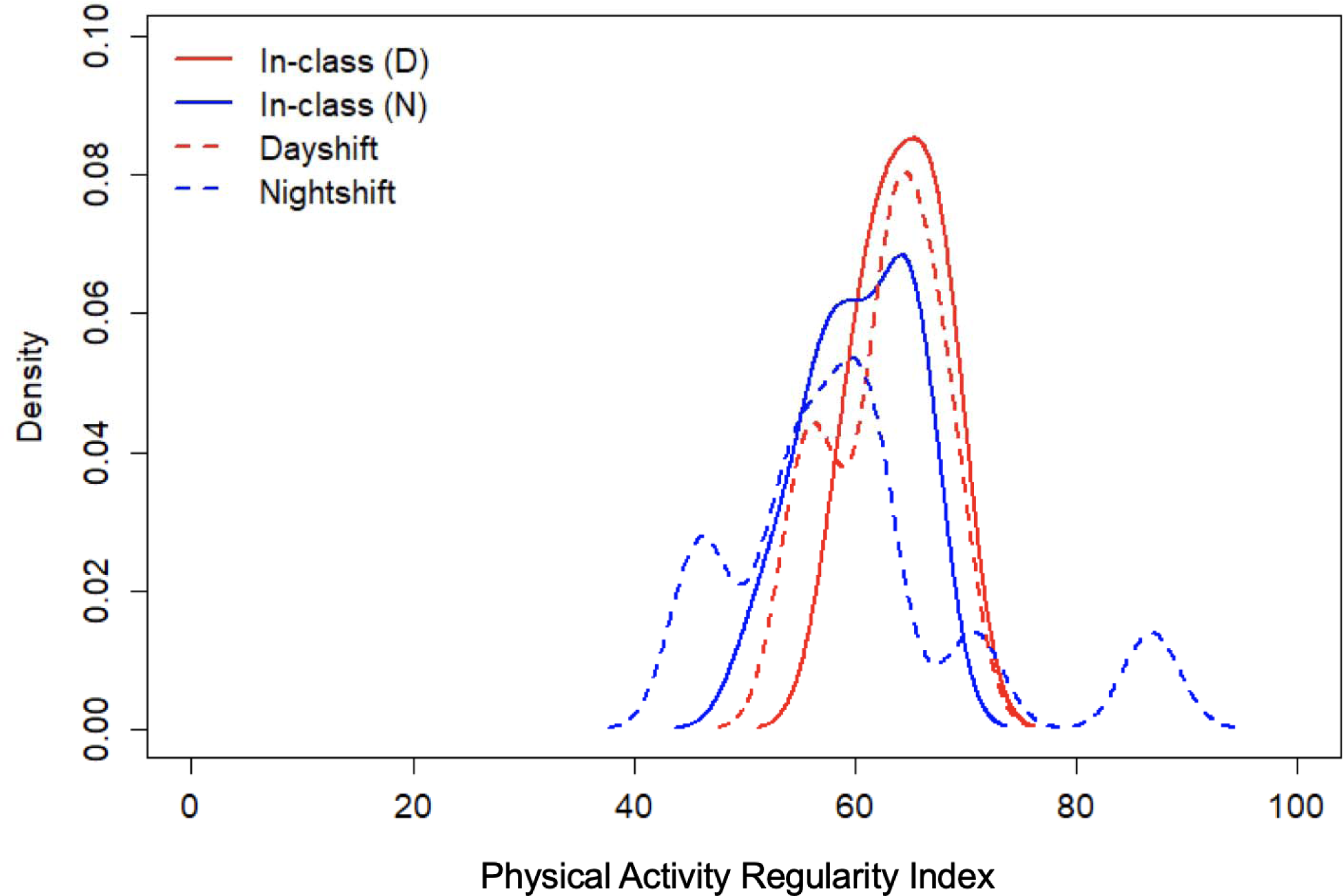
Regularity of Physical Activity Timing is Similar on Dayshift and Nightshift. Figure shows density plots of Physical Activity Regularity Index (PARI) of the participant cohort during the transition from dayshift in-class training (In-class (D); red line) to dayshift field-training (Dayshift; dashed red line), as well as during the transition from dayshift in-class training (In-class (N); blue line) to nightshift field-training (Nightshift; dashed blue line). Each index is expressed as a value ranging from 0 to 100, in which 0 reflects irregularity and 100 reflects regularity.

#### Meal Timing Regularity Index

The transition from dayshift in-class training to dayshift field-based training was not significantly different (84.5 vs. 84.1, *P*=0.375; **Table 1**). In contrast, the transition from dayshift in-class training to nightshift field-based training led to a significant reduction in meal timing regularity (85.5 vs. 49.2, *P*=0.001; **Table 1**). Moreover, changes in the MRI were significantly different between groups (*P*<0.001; Table 1). No differences were observed between groups during the dayshift in-class training (*P*=0.928). **Figure 4** shows density plots of the MRI during all four conditions.

**Figure 4:**
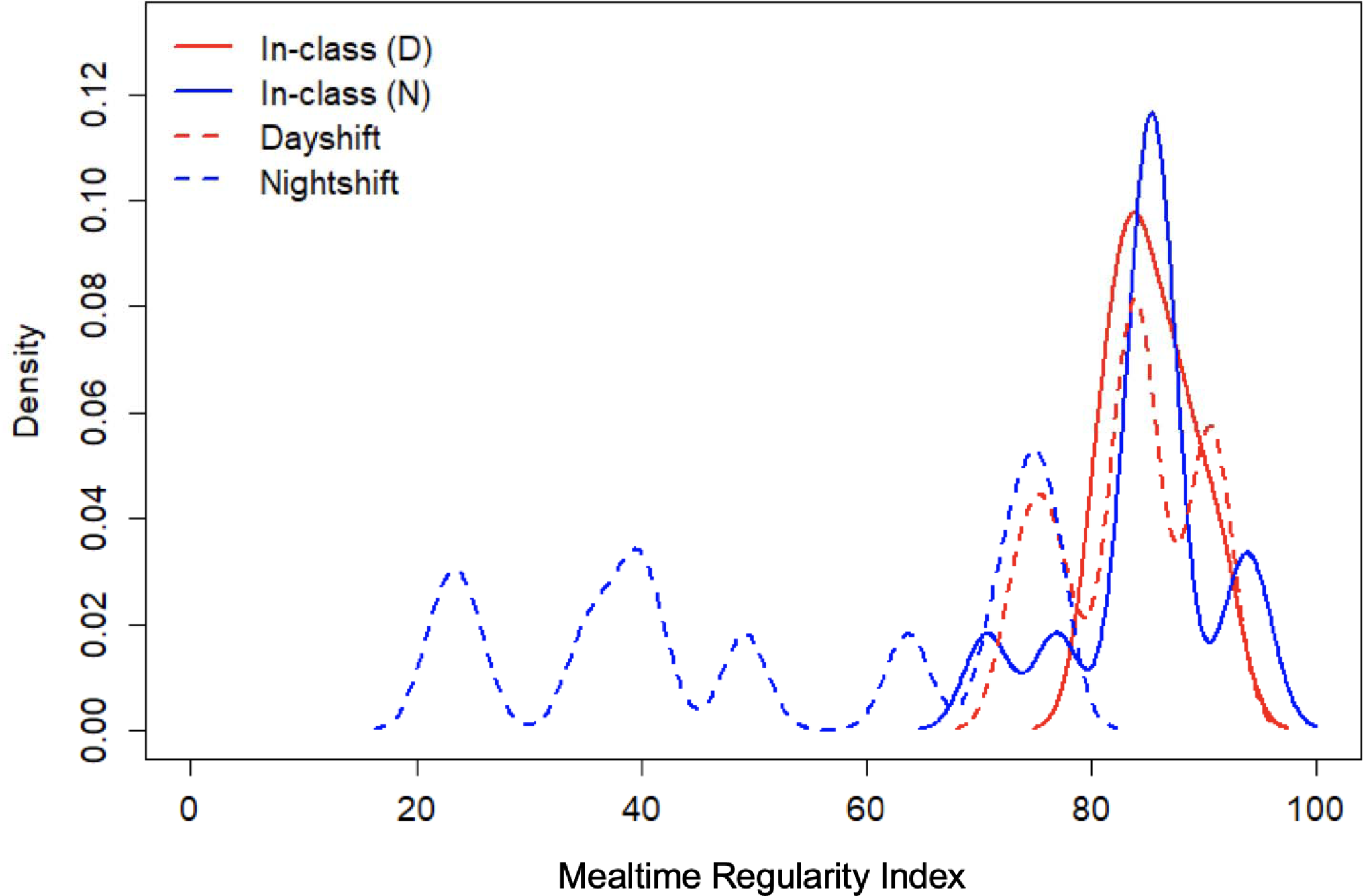
Regularity of Meal Timing is Reduced on Nightshift. Figure shows density plots of Mealtime Regularity Index (MRI) of the participant cohort during the transition from dayshift in-class training (In-class (D); red line) to dayshift field-training (Dayshift; dashed red line), as well as during the transition from dayshift in-class training (In-class (N); blue line) to nightshift field-training (Nightshift; dashed blue line). Each index is expressed as a value ranging from 0 to 100, in which 0 reflects irregularity and 100 reflects regularity.

### Effect of Shiftwork on Behavior Regularity Index

The transition from dayshift in-class training to dayshift field-based training on the composite BRI was not significantly different in dayshift (1.00 vs. 1.00; *P*=0.250; **Table 1**). In contrast, the transition from dayshift in-class training to nightshift field-based training reduced composite BRI (1.00 vs. 0.00, *P*=0.001; **Table 1**). Moreover, changes in the BRI were significantly different between dayshift and nightshift groups (*P*<0.001; **Table 1**). No differences were observed between groups during dayshift in-class training (*P*=0.359). **Figure 5** shows density plots of the BRI during the transition from dayshift in-class training to dayshift field-based training (**Panel A**) and dayshift in-class training to nightshift field-based training (**Panel B**).

**Figure 5:**
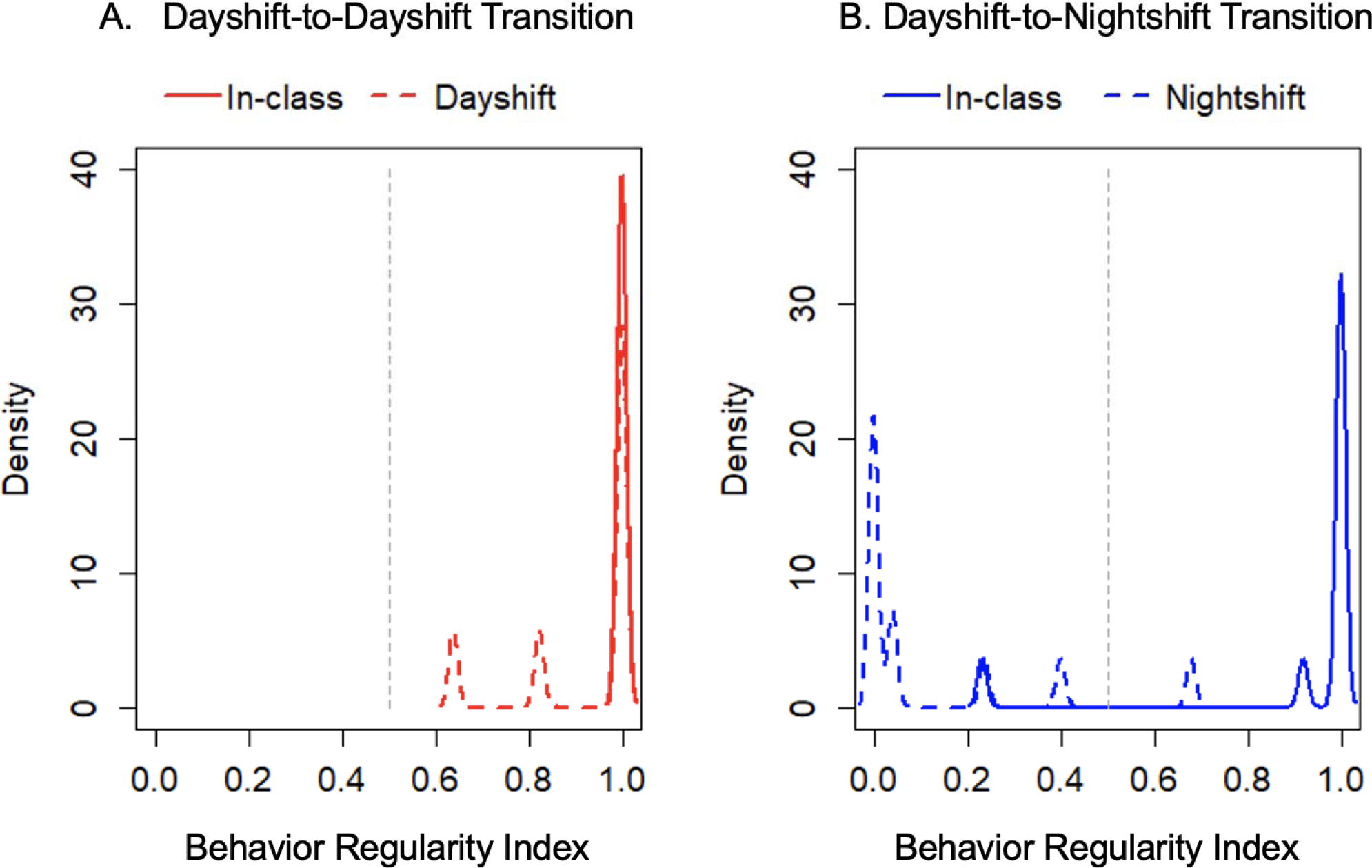
Regularity of Lifestyle Behavior Timing is Reduced on Nightshift. **Panel A** shows density plots of BRI of the participant cohort during the transition from dayshift in-class training (In-class; red line) to dayshift field-training (Dayshift; dashed red line). **Panel B** shows density plots of BRI from participant cohort during the transition from dayshift in-class training (In-class; blue line) to nightshift field-training (Nightshift; dashed blue line). BRI is expressed as a value ranging from 0 to 1, in which 0 reflects maximum irregularity and 1 reflects maximum regularity. The dashed grey line marks a reference point of 0.5, in which a BRI between 0.5 and 1 is classified as ‘regular’ and a BRI between 0 and 0.49 is classified as ‘irregular’.

The BRI was calculated and rounded to the nearest integer to classify participant behaviors as either regular or irregular. Our *a priori* hypothesis was that dayshift schedules would produce a BRI classified as regular (closer to 0.5-1) and nightshift schedules would produce a BRI classified as irregular (closer to 0-0.5). As shown in the contingency table in **Table 2**, this hypothesis was confirmed for 24 of the 25 participant dayshift observations classified as regular and 10 of 11 participant nightshift observations classified as irregular. The odds ratio for each behavior component index was calculated, but not significant (**Table 3**). Of the three indices, though, the MRI had the largest odds ratio (2.16) compared to the PARI (1.39) and SRI (1.09).

**Table 2:** Comparison of Predicted Behavior Regularity Index vs. Actual Shiftwork Schedule

| **Computed Behavior Regularity** | **Actual Shiftwork Schedule** |  |
| --- | --- | --- |
|  | *Dayshift* | *Nightshift* |
| *Regular* | 24 | 1 |
| *Irregular* | 1 | 10 |

Computed behavior regularity index was calculated using linear regression. We then compared the predicted shiftwork schedule (e.g., regular behavior index was hypothesized to indicate an actual dayshift schedule, whereas an irregular behavior index was hypothesized to indicate an actual nightshift schedule).

**Table 3:** Odds Ratio of Component Input Indices

| **Parameter** | **Odds Ratio** | **95% CI** | ***P*-value** |
| --- | --- | --- | --- |
| Sleep Regularity Index | 1.09 | (0.89, 1.34) | 0.401 |
| Physical Activity Regularity Index | 1.39 | (0.86, 2.25) | 0.184 |
| Mealtime Regularity Index | 2.16 | (0.82, 5.68) | 0.119 |

## DISCUSSION

The primary finding of this study was that nightshift reduced the regularity of lifestyle behavior timing, whereas dayshift enabled maintenance of lifestyle behavior regularity. This finding is consistent with previous work in shift-working nurses that, through the use of self-report logs, indicate a variety of sleep/wake patterns are used to cope with shiftwork demands ^20^. We extended this earlier work to also consider two additional behaviors that are relevant for health, namely physical activity and meal timing. This current work addresses a knowledge gap by generating empirical data to confirm that sleep/wake patterns and meal timing are irregular during nightshift in the real-world. Controlled in-patient studies reveal that following a simulated nightshift schedule has acute detrimental consequences on cardiometabolic health ^10–12^. Our findings of irregular sleep/wake patterns and meals during nightshift in sample of police trainees—for which shiftwork is unavoidable—supports the translation and relevance of inpatient studies to real-world settings of shiftwork characterized by repeated bouts of nightshift.

The novel BRI quantifies the degree of regularity with which behavioral events occur at the same point in time, 24 hours apart, on a day-to-day timescale. As a test of internal validity, we compared BRI against the known shiftwork schedule, with the *a priori* expectation that participants on nightshift would exhibit reduced behavior regularity, whereas participants on dayshift would exhibit maintenance of behavior regularity. On average, we had successful predictions for matching of behavior regularity compared to the shiftwork schedule. Specifically, 91% of the observations from nightshift were classified as irregular and 96% of the observations from dayshift were classified as regular. Indeed, this analysis revealed some interesting misclassifications. We observed that one participant on nightshift exhibited regularity in lifestyle behavior patterns by maintaining a ‘nightshift’ schedule during non-workdays (**Figure 6; Panel A**). Additionally, we observed that one participant exhibited irregularity in lifestyle behavior patterns during in-class training (**Figure 6; Panel B**), possibly related to illness that altered sleep/wake patterns and induced irregular mealtimes (MRI=70). Overall, these findings suggest that, on average, nightshift disrupts the regularity in lifestyle behaviors from day-to-day, whereas dayshift enables a maintenance of behavior regularity.

**Figure 6:**
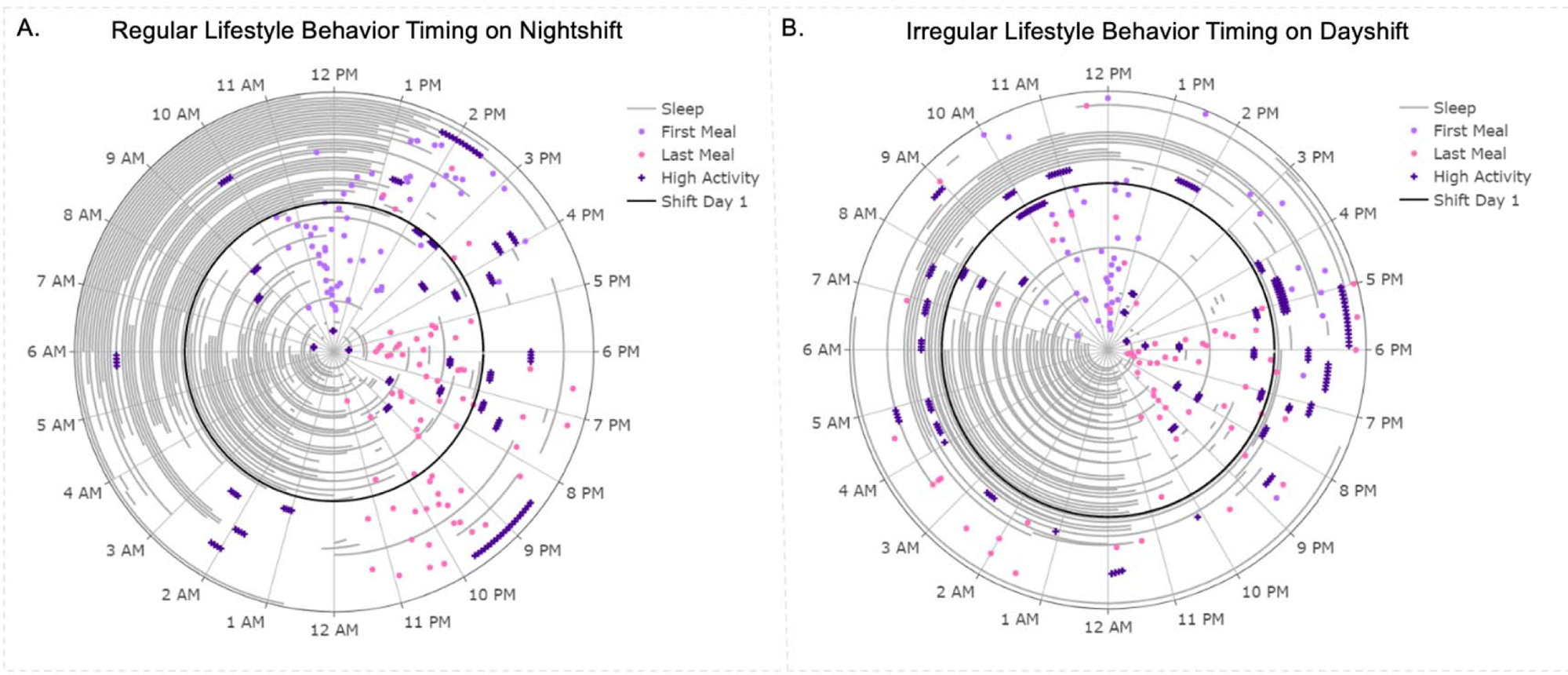
Regular Lifestyle Behaviors on Nightshift and Irregular Lifestyle Behaviors on Dayshift. Representative polar plots for two instances in which BRI did not correspond with actual shiftwork schedule. **Panel A** shows an example of behavior regularity during the 6 weeks of nightshift, indicating that a nightshift schedule was regularity maintained during non-workdays. **Panel B** shows an example of behavior irregularity during 6 weeks of in-class training, despite a typical dayshift schedule.

We primarily focused on regularity of three lifestyle behaviors, which included sleep/wake patterns, physical activity, and meals. We also calculated the odds ratio which describes the increase in likelihood that a participant will be classified as regular for a 1-point increase in a component index. Of these three behaviors, meal timing had the largest odds ratio (2.16) relative to the physical activity (1.39) and sleep (1.09), suggesting that this behavior had the strongest influence on the BRI, although not statistically significant. The observation that nightshift led to irregular meal timing may be concerning in the context of weight gain; late meal timing relative to circadian phase is associated with increased adiposity ^21, 22^. Nightshift also led to irregular sleep/wake patterns, in agreement with previous work ^20^. Physical activity regularity was minimally impacted by nightshift, and this finding may be explained by the low levels of physical activity in our participant cohort. Given the well accepted health benefits of physical activity and exercise, this finding may highlight a targetable behavior for intervention specific to shift workers. Thus, an advantage of the BRI is it is designed to be sensitive to changes in behavioral patterns in physical activity, sleep/wake, and meal timing, over a broad range of adaptations in a variety of populations.

As with any study utilizing field-based assessments, we encountered data missingness regarding self-reported mealtimes and sleep/wake patterns due to non-compliance. In addition, there may be some misclassification in sleep/wake patterns – specifically when the sleep detection algorithm noted sleep periods while the subject was supposedly in the classroom or on patrol due to low activity levels and heart rate. In these situations, classification of meals relative to sleep was done manually and subjectively, considering the participants’ behavior patterns on surrounding days when both meal and activity data were collected. Meals reported during sleep were not accounted for in the analysis, nor were meals occurring between meal 1 and meal 3 since not all participants consumed 3 meals a day. Further, with respect to the length of fasting periods, the intermediate meals likely are less physiologically important. Lastly, any meal that was the only meal reported for a given waking period was regarded as both meal 1 and meal 3.

In summary, we observed that nightshift imposed a rapid and sustained reduction in regularity of lifestyle behavior timing as compared to dayshift. We also identified ‘behavioral’ outliers, in which the anticipated behavior regularity did not necessarily match with the shiftwork schedule, suggesting some variation in behavioral strategies to cope with shiftwork. Through the use of wearable activity trackers and mobile devices, we demonstrate the use of a novel BRI metric that quantifies lifestyle behavior regularity in real-world settings of shiftwork. One advantage of this approach is that it is feasible to scale these assessments to larger participant cohorts across long time-domains. Future studies might consider using the BRI to gain new insight into patterns of behavior in other populations experiencing circadian disruption.

## Data Availability

All data produced in the present work are contained in the manuscript

## ACKWOLEDGEMENTS

Authors would like to thank the Greensboro Police Department for participants in this study. This study was supported by 1P30 AG028716, Claude D Pepper Older Americans Independence Centers (OAICs) to Duke University. MLE supported in part by K01DK134838.

